# Examining Medical Student Volunteering During The COVID-19 Pandemic As A Prosocial Behavior During An Emergency

**DOI:** 10.1101/2021.07.06.21260058

**Authors:** Matthew H V Byrne, James Ashcroft, Jonathan C M Wan, Laith Alexander, Anna Harvey, Nicholas Schindler, Megan E L Brown, Cecilia Brassett

## Abstract

**Introduction:** COVID-19 has caused major disruptions to healthcare, with voluntary opportunities offered to medical students to provide clinical support. We used the conceptual framework of prosocial behavior during an emergency – behaviors whose primary focus is benefiting others – to examine volunteering during COVID-19.

**Methods:** We conducted an in-depth, mixed-methods cross-sectional survey, from 2^nd^ May to 15^th^ June 2020, of medical students studying at UK medical schools. Data analysis was informed by Latane and Darley’s theory of prosocial behavior during an emergency and aimed to understand students’ decision-making processes.

**Results:** A total of 1145 medical students from 36 medical schools completed the survey. While 947 (82.7%) of students were willing to volunteer, only 391 (34.3%) had volunteered. The majority (92.7%) of students understood that they may be asked to volunteer; however, we found that deciding one’s responsibility to volunteer was mitigated by a complex interaction between the interests of others and self-interest. Further, concerns revolving around professional role boundaries influenced students’ decisions over whether they had the required skills and knowledge to volunteer. Deciding to volunteer depended not only on possession of necessary skills, but also seniority and identification with the nature of volunteering roles offered.

**Conclusions:** We propose two additional domains to Latane and Darley’s theory of prosocial behavior during an emergency that students consider before making their final decision to volunteer. These are ‘logistics’ – whether it is logistically feasible to volunteer – and ‘safety’ – whether it is safe to volunteer. This study highlights a number of modifiable barriers to prosocial behavior that medical students encounter and provides suggestions regarding how Latane and Darley’s theory of prosocial behavior can be operationalized within educational strategies to address these barriers. Optimizing the process of volunteering can aid healthcare provision and may facilitate a safer volunteering process for all.

## INTRODUCTION

On 24^th^ March 2020, the UK Secretary of State for Health and Social Care announced plans for medical students to assist in the COVID-19 pandemic ^1^. In response to this, medical schools accelerated the graduation of over 5000 final year medical students to act as interim postgraduate year one doctors ^2^, and volunteering opportunities were created for non-final year students ^3^. As clinical placements were cancelled and medical school examinations postponed or replaced, there was the potential for many students to volunteer ^4,5^.

Volunteering is a form of prosocial behavior – behavior which provides help to others where a direct personal benefit is not a necessity but self-interest is considered ^6,7^. As such it can have positive benefits for both doctors and patients ^7^. Prosocial behavior is an important professional value in medicine ^8,9^. Indeed, the Situational Judgement Test that final year medical students in the UK must pass prior to graduation examines prosocial decision making^10^. McCrea and Murdoch-Eaton highlighted that while medical students recognize the role of prosocial behavior for doctors, they often possess limited awareness of the role of prosocial behavior within their current role as students ^11^. This has relevance for student volunteering during COVID-19 – a form of prosocial behavior during an emergency. Latane and Darley proposed that, in prosocial behavior theories, five factors influence an individual’s decision to help during an emergency ^12^, which Baron et al. posited to be the same for volunteering ^6^. They are: 1. Noticing something is abnormal; 2. Interpreting the situation as an emergency; 3. Deciding you are responsible; 4. Deciding whether you have the skills or knowledge to help; and 5. Making a final decision on providing help ^6,12^. It has yet to be established whether Latane and Darley’s theory of prosocial behavior applies to medical student volunteering during COVID-19.

Studies prior to the COVID-19 pandemic have reported the readiness of medical students to volunteer in hypothetical disasters and infectious disease outbreaks ^13,14^, and early studies into COVID-19 indicate that the majority of students appear willing to volunteer ^15,16^. Rasmussen et al. showed that a high percentage of medical students from a single centre in Denmark wanted to volunteer (82.4%) ^15^. However, other studies have shown that, whilst students are willing to volunteer during COVID-19, fewer students have actually done so ^17–19^. A single centre study of 137 German medical students demonstrated that 70.1% were willing to volunteer, but only 25.0% of students did ^18^. Despite this discrepancy between motivation and practice, minimal literature exists on the factors motivating students to volunteer during COVID-19 ^18,20,21^. Understanding the factors that influence prosocial behavior during the current pandemic is essential, as over 40,000 medical students who are studying in the UK could represent a valuable asset if empowered and mobilized as volunteers ^22^.

In view of the above, we conducted an in-depth, mixed-methods survey to explore volunteering among UK medical students during COVID-19, using the conceptual framework of prosocial behavior described by Latane and Darley, and Baron et al. ^6,12^. Through developing an understanding of students’ motivations to volunteer, we aimed to identify educational strategies which support prosocial behaviors during emergencies to support medical education volunteering pathways.

## METHODS

### Research approach

This research was conducted within the paradigm of pragmatism. Pragmatism concerns itself with problem solving, and often utilizes mixed-methods approaches ^23^. In line with our pragmatic orientation, we chose not to forefront considerations of epistemology and ontology ^24^, and focused on designing an effective study expeditiously within the new research landscape mandated by COVID-19. As such, an online survey was selected for data collection for ease of dissemination and wide reach. Questions were asked with both quantitative and qualitative outputs to provide broad and rich data reflective of a wide range of experiences ^25^.

### Survey

We conducted a cross-sectional survey from 2^nd^ May to 15^th^ June 2020 of students studying at UK medical schools, following the STROBE guideline for cross-sectional studies ^26^. The survey consisted of 53 questions assessing previous clinical experience, attitudes to volunteering, motivation and barriers, volunteering role, medical education, issues currently faced, and safety (Appendix 1). Survey development was informed by a systematic review of existing literature on volunteering during pandemics and disasters, and previously used scales^27^. Questions were then developed by MHVB and JA with expert input and consultation with medical students, and final questions were reviewed by medical students to establish face validity. The survey was hosted on Google Forms with no identifiable data collected, and data were held on a secure server.

### Student recruitment

We used a convenience sampling approach to recruit medical students. Medical schools listed on the UK Medical Schools Council website were invited via email to distribute the survey to their students ^28^. Messages were posted once a week to Twitter and Facebook asking medical students to complete and share the survey to recruit more participants via a snowball approach.

### Ethics

Ethical approval for the study was obtained from the University of Cambridge Psychology Research Ethics Committee (PRE.2020.040).

### Quantitative analysis

We asked students to indicate their level of agreement using a 5-point Likert scale (1=strongly disagree; 5=strongly agree) on items representing volunteering, role, clinical skills, motivation/barriers to volunteering, issues with volunteering, and risk and safety. Statistical analysis was performed by JW using R (version 4.0.1) ^29^. Data are presented as mean ± standard deviation (SD) and as percentages in each category of the Likert scale. Correction for multiple testing was performed using a Benjamini-Hochberg correction in R using the rstatix package.

Multiple linear regression of predictors for ‘I am willing to volunteer (Likert)’ was performed in R using lm(). For this model, the variables used were ‘year at medical school’ and survey responses around beliefs for the prediction of volunteering status. Data for subgroups were analysed separately and included in the multiple regression model. These groups were chosen to ascertain whether attitudes differed between years at medical school and if any differences existed between those who chose to volunteer and those who did not. Entries with missing data were excluded from the analysis using the na.omit function in R. Backward stepwise model selection was performed using stepAIC() from the MASS package.

### Qualitative analysis

Thematic analysis was used to analyse qualitative responses using the six-step approach described by Braun and Clarke ^30^. Two authors (LA and MHVB) familiarised themselves with the data, and created initial inductive, descriptive codes for all data. To identify themes, a semantic approach was used. Initial codes were analysed for patterns, grouped, summarised, and interpreted. Themes and subthemes were checked against the initial codes and the data set as a whole, and any interpretative discrepancies explored and resolved by consensus. Themes and subthemes were then reviewed, discussed, and agreed by all authors.

### Applying Latane and Darley’s theory of prosocial behavior during an emergency as a theoretical lens

To analyse how our data applied to Latane and Darley’s theory of prosocial behavior during an emergency, we used a theory-informing inductive data analysis approach ^31^. We did not impose our definition of prosocial behavior during an emergency upon participants, instead allowing them to express their own views in response to questions. We first performed our quantitative and qualitative analysis, later applying Latane and Darley’s theory to our data as a ‘sensitizing concept’. Though sensitizing concepts originate from the methodology of constructivist grounded theory ^32^, they have since been applied as part of reflexive analysis in a way that aligns with Varpio et al.’s ‘theory informing inductive data analysis’ approach ^33^. This is the way in which we utilized theory within this study.

Data were reviewed using this theoretical framework by MHVB and MELB, and areas of concordance and conflict explored and highlighted, then discussed and agreed by all authors.

### Reflexivity statement

The authors of this study comprise a diverse range of doctors in training, medical students, medical education researchers, and consultants across multiple educational institutions. The range of experiences at different stages and institutions permitted a wide scope of viewpoints regarding the data, thus enriching analysis.

## RESULTS

### Demographics

A total of 1145 students from 36 medical schools were represented in this study (Figure S 3: *Roles students are willing to perform while volunteering grouped by year*.). The median age of respondents was 22 (interquartile range, IQR, 20-24), 835 (73.0%) were women, 75 (6.6%) were intercalating (taking time out of medical school to complete an additional degree), and 170 (14.8%) were in their fifth or sixth year of medical school (Figure S 1). Of these final-year students, 112 out of 170 (65.9%) had already graduated.

### Quantitative analysis

Results are structured within both qualitative and quantitative sections following the domains of Latane and Darley’s theory of prosocial behavior during an emergency.

#### Noticing something is abnormal and interpreting the situation as an emergency

The majority of students (92.7%) recognized that medical students might be asked to volunteer due to COVID-19. We interpret this as widespread recognition of an abnormal, emergency situation.

#### Deciding responsibility

Across all year groups, 947 out of 1145 students (82.7%) strongly agreed or agreed with the statement ‘I would be willing to volunteer to work’, which suggests that most students felt responsible for helping (Figure S 2). The reasons for this were complex, with a wide range of motivating and barrier factors for volunteering (Table 1). Factors influencing the responsibility to volunteer can be divided into two groups: in the interest of others and self-interest. The former included altruism, moral obligation, family or social commitments, others’ safety, societal expectations, medical school expectations, and peer pressure; the latter included professional development, academic and work commitments, personal safety, psychological impact, discrimination, and financial implications.

**Table 1:**
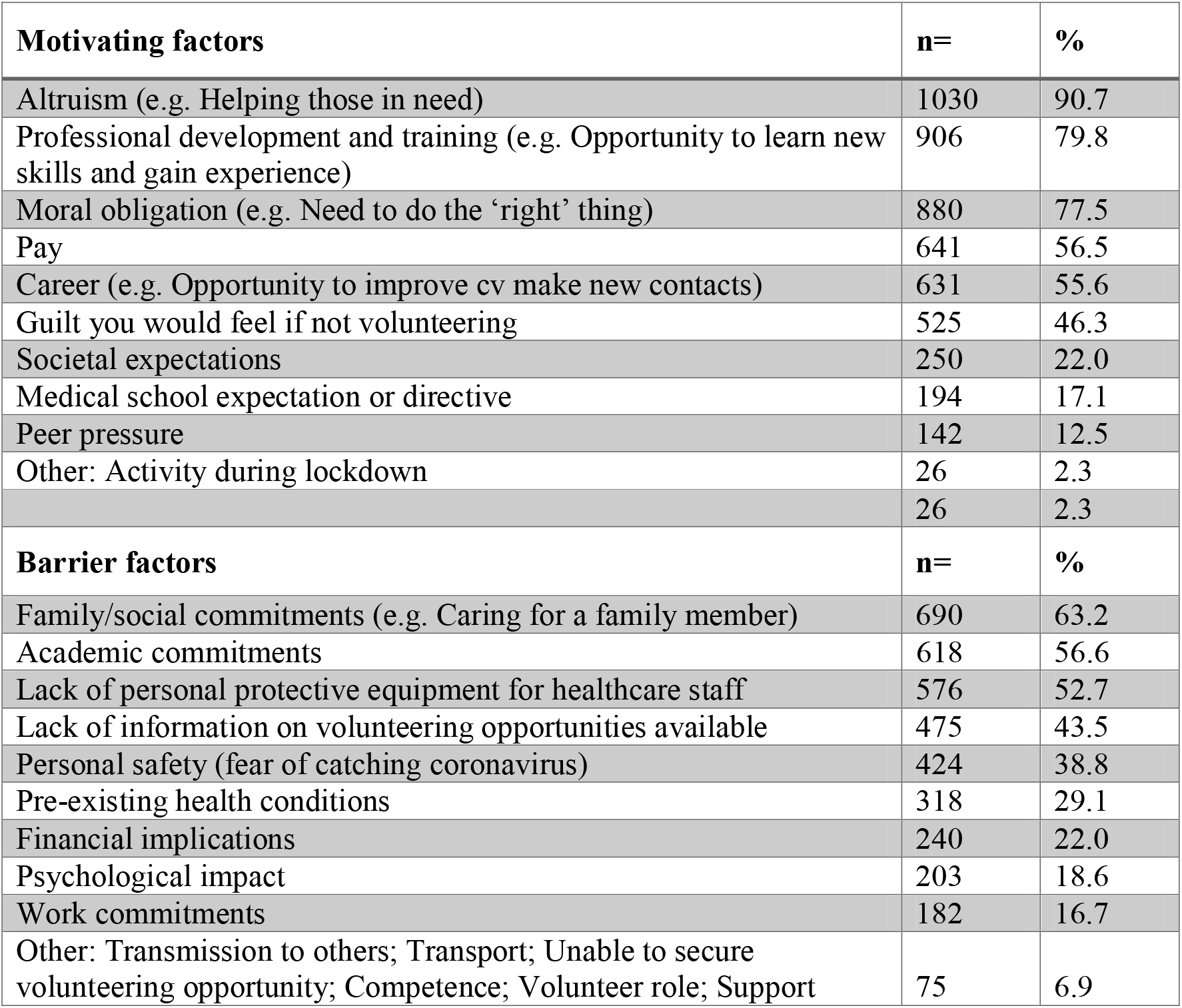
Motivating and barrier factors for volunteering

While most students were willing to volunteer, less than half of the students felt they should be encouraged to volunteer (Figure S 2).

#### Deciding whether you have the skills or knowledge to help

Eighty percent of students felt they would have a positive impact by volunteering. We assessed students’ preference for specific volunteer roles, and their confidence in the skills required for these roles.

We asked students to indicate the roles they were willing to perform as volunteers. Twenty-six percent of students strongly agreed or agreed that they were willing to perform the full clinical role of a doctor, whereas 857 (75.3%) and 882 (77.7%) were willing to undertake an assistant medical role or provide indirect medical care (such as providing meals or moving patients), respectively. The majority of students were willing to perform the same role on a ward with patients with COVID-19 (n=943, 82.3% strongly agreed or agreed, Figure 1). Senior students were more willing to perform the full clinical role expected of a doctor but were less willing to provide indirect medical care (Figure S 3).

**Figure 1:**
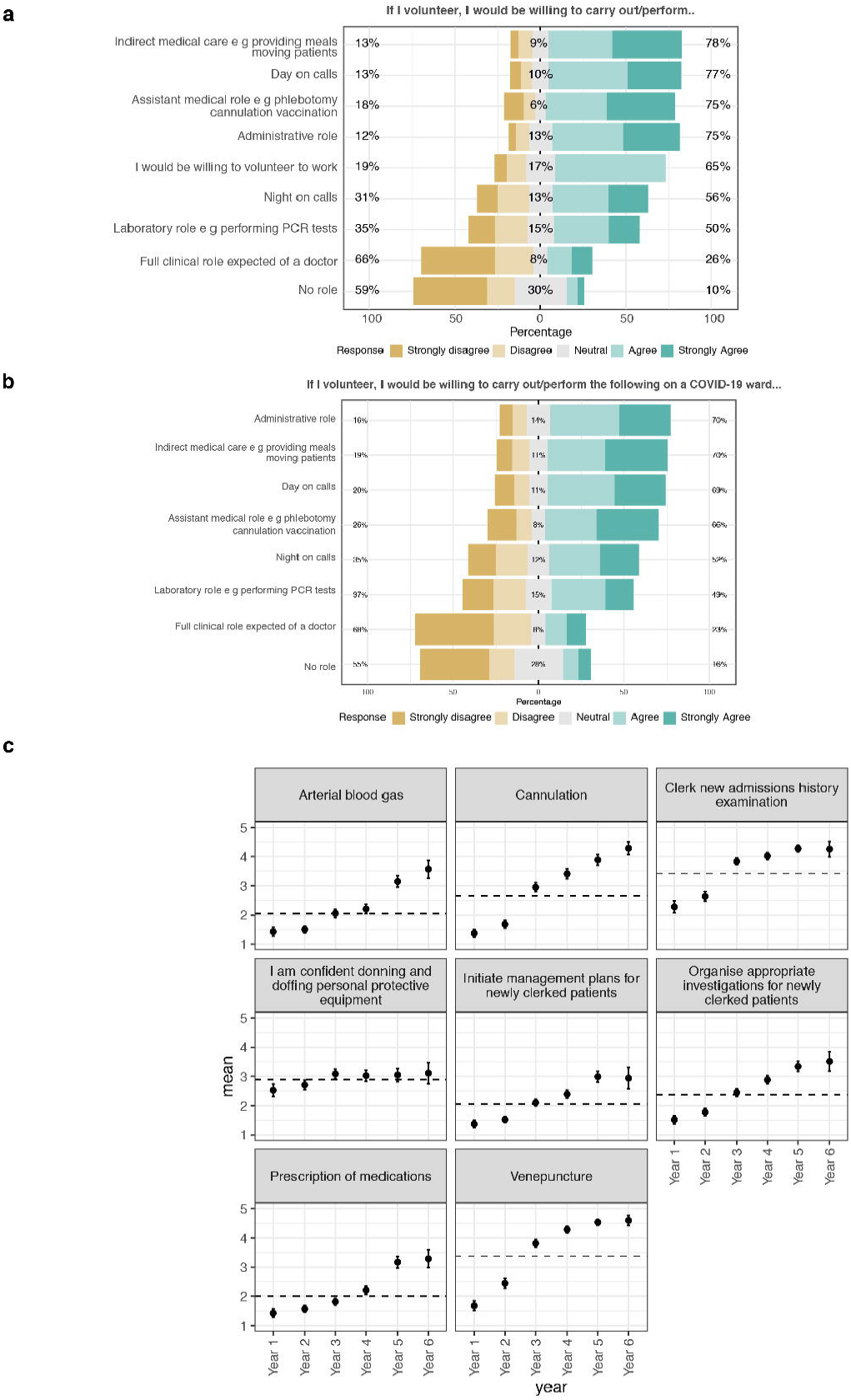
*Willingness to perform roles during the pandemic, both (a) on a general ward, and (b) on a coronavirus ward. (c)* Confidence in clinical skills by year. Mean values and 95% confidence intervals are shown. The mean confidence score across all students is shown with a dashed line.

We looked at student confidence in skills required for these roles using a Likert scale (Figure 1). Across all year groups, students were most confident in clerking new admissions (mean=3.4/5, 95% CI=3.3-3.5) and performing venepuncture (mean=3.4/5, 95% CI=3.3-3.5). Students were least confident in prescribing medication (mean=2.0/5, 95% CI=1.9-2.0) and initiating management plans for patients (mean=2.1, 95% CI=2.0-2.1). For all skills except for donning and doffing PPE, there was a significant positive correlation between the respondents’ year group and their confidence in performing them (Benjamini-Hochberg adjusted P<0.05).

#### Making a final decision to provide help by volunteering

At the time of the study, 391 of 1145 students (34.3%) had volunteered during the pandemic (Figure S 1). This ranged from 14.6% amongst first-year students to 54.1% in final year (year five or six). A significantly lower proportion of intercalating students were volunteering compared to their clinical counterparts in the same years (20.0% vs. 33.5%, P=0.02, Chi-square test). The median start date of those who had begun volunteering was 16^th^ April 2020, three weeks after the UK was placed into lockdown. Of the 391 students who were volunteering in the pandemic, 77 (22.2%) had taken up roles as interim postgraduate year one doctors (Table S 1).

The strongest predictors of willingness to volunteer using multiple linear regression were the beliefs that volunteering to work would benefit a student’s medical education (estimate=0.35±0.03, adjusted P<0.001, Table S 2) and that the student would make a positive impact (estimate=0.33±0.03, adjusted P<0.001). Students who believed there were ethical issues with asking medical students to volunteer were less likely to volunteer (estimate=-0.08±0.02, adjusted P<0.001), as were those who had begun considering a career outside of medicine because of the pandemic (estimate=-0.08±0.02, adjusted P=0.001). Increasing age was a significant negative predictor of willingness to volunteer, independent of medical school year group, after correction for multiple testing (adjusted P=0.043).

#### Additional factors which influenced making a final decision to provide help

Across all year groups, the median self-estimated probability of contracting COVID-19 was 50.0% (IQR = 20.0%-65.0%, n = 962), which was not influenced by year or volunteering. 43.5% (n=475) felt there was a lack of information regarding volunteering opportunities available.

### Qualitative analysis

Our qualitative analysis focused on the later stages of the decision-making process. We asked students what ethical concerns they had about volunteering and the issues they anticipated they would face while volunteering (Table 2). We defined five themes: Pressure to volunteer; Education; Professional practice; Safety; and Logistics.

**Table 2:**
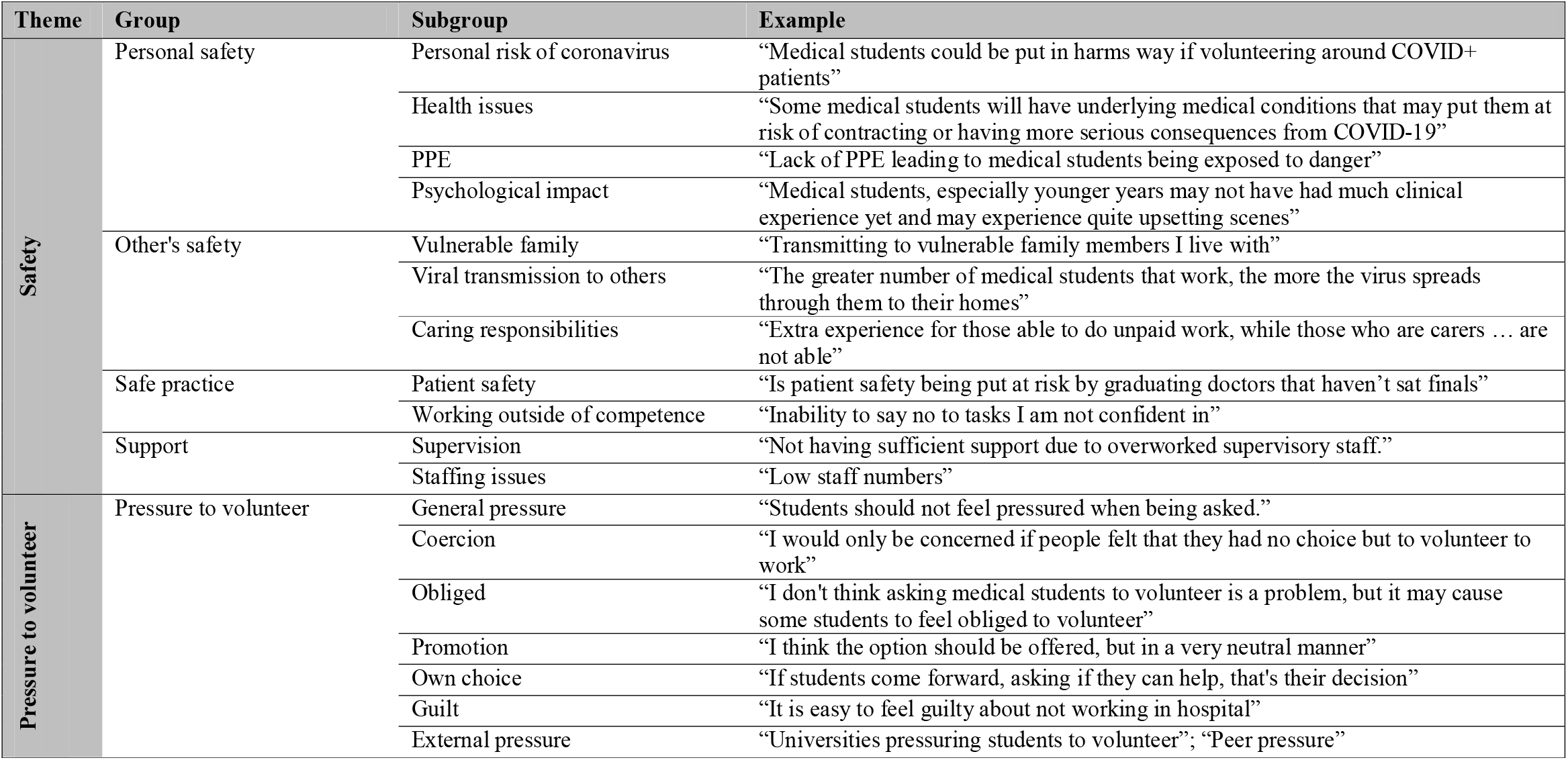
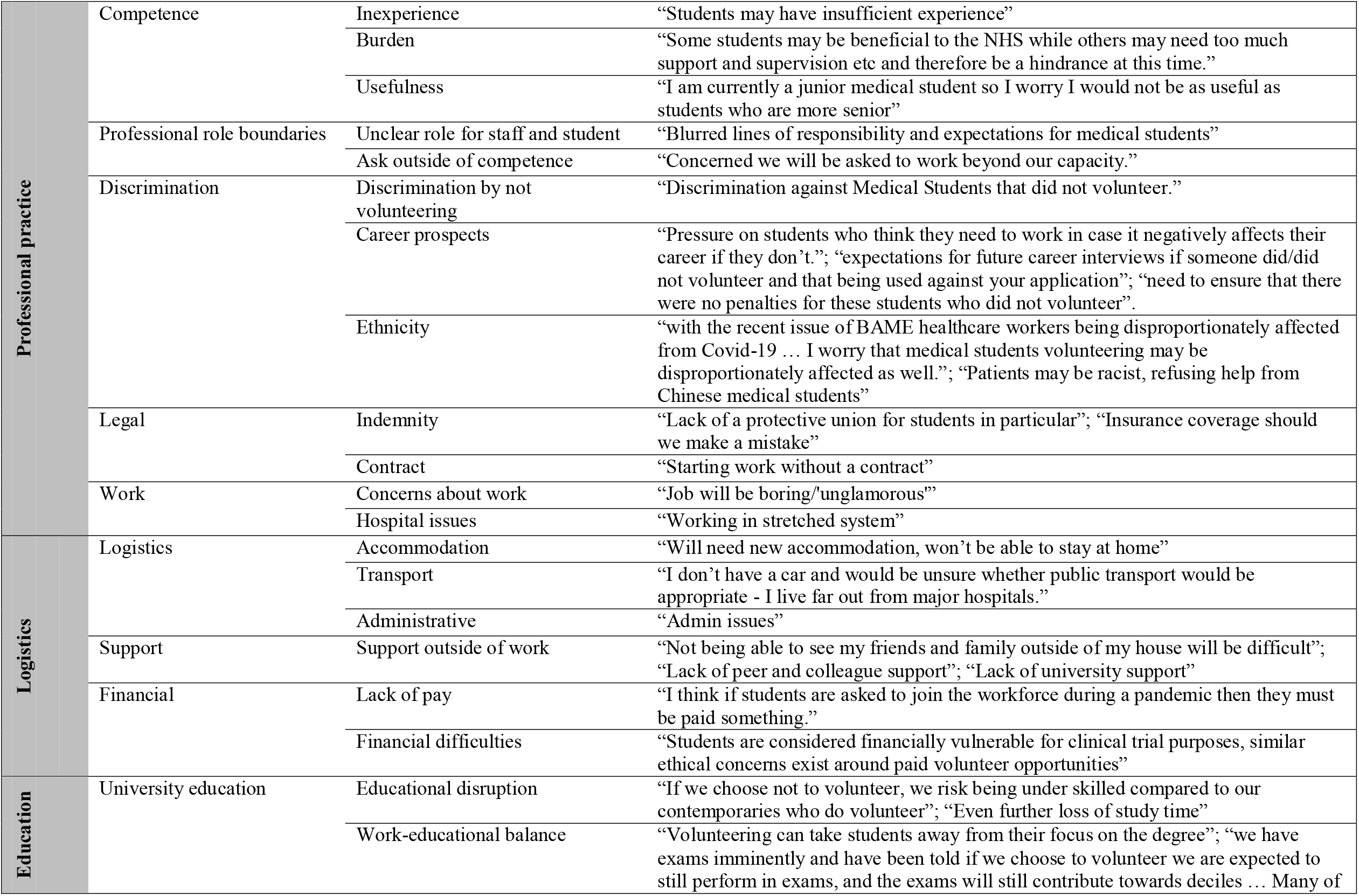
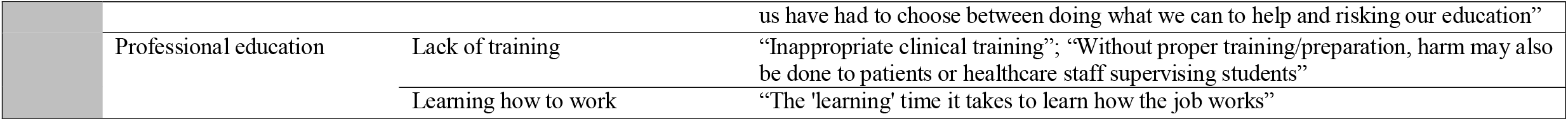
Concerns about volunteering during a pandemic from medical students. 798 out of 1145 students (69.7%) described issues they believed they would face when volunteering, and 25 students stated they predicted no issues. 365 out of 1145 students (31.9%) described ethical concerns associated with volunteering, and three students stated there were no ethical concerns. From these responses we identified five themes: safety, professional practice, pressure to volunteer, finances and logistics, and education.

| Theme | Group | Subgroup | Example |
| --- | --- | --- | --- |
| Safety | Personal safety | Personal risk of coronavirus | “Medical students could be put in harms way if volunteering around COVID+ patients” |
|  |  | Health issues | “Some medical students will have underlying medical conditions that may put them at risk of contracting or having more serious consequences from COVID-19” |
|  |  | PPE | “Lack of PPE leading to medical students being exposed to danger” |
|  |  | Psychological impact | “Medical students, especially younger years may not have had much clinical experience yet and may experience quite upsetting scenes” |
|  | Other's safety | Vulnerable family | “Transmitting to vulnerable family members I live with” |
|  |  | Viral transmission to others | “The greater number of medical students that work, the more the virus spreads through them to their homes” |
|  |  | Caring responsibilities | “Extra experience for those able to do unpaid work, while those who are carers ... are not able” |
|  | Safe practice | Patient safety | “Is patient safety being put at risk by graduating doctors that haven't sat finals” |
|  |  | Working outside of competence | “Inability to say no to tasks I am not confident in” |
|  | Support | Supervision | “Not having sufficient support due to overworked supervisory staff.” |
|  |  | Staffing issues | “Low staff numbers” |
| Pressure to volunteer | Pressure to volunteer | General pressure | “Students should not feel pressured when being asked.” |
|  |  | Coercion | “I would only be concerned if people felt that they had no choice but to volunteer to work” |
|  |  | Obligated | “I don't think asking medical students to volunteer is a problem, but it may cause some students to feel obliged to volunteer” |
|  |  | Promotion | “I think the option should be offered, but in a very neutral manner” |
|  |  | Own choice | “If students come forward, asking if they can help, that's their decision” |
|  |  | Guilt | “It is easy to feel guilty about not working in hospital” |
|  |  | External pressure | “Universities pressuring students to volunteer”; “Peer pressure” |
| Professional practice | Competence | Inexperience | “Students may have insufficient experience” |
|  |  | Burden | “Some students may be beneficial to the NHS while others may need too much support and supervision etc and therefore be a hindrance at this time.” |
|  |  | Usefulness | “I am currently a junior medical student so I worry I would not be as useful as students who are more senior” |
|  | Professional role boundaries | Unclear role for staff and student | “Blurred lines of responsibility and expectations for medical students” |
|  |  | Ask outside of competence | “Concerned we will be asked to work beyond our capacity.” |
|  | Discrimination | Discrimination by not volunteering | “Discrimination against Medical Students that did not volunteer.” |
|  |  | Career prospects | “Pressure on students who think they need to work in case it negatively affects their career if they don’t.”; “expectations for future career interviews if someone did/did not volunteer and that being used against your application”; “need to ensure that there were no penalties for these students who did not volunteer”. |
|  |  | Ethnicity | “with the recent issue of BAME healthcare workers being disproportionately affected from Covid-19 ... I worry that medical students volunteering may be disproportionately affected as well.”; “Patients may be racist, refusing help from Chinese medical students” |
|  | Legal | Indemnity | “Lack of a protective union for students in particular”; “Insurance coverage should we make a mistake” |
|  |  | Contract | “Starting work without a contract” |
|  | Work | Concerns about work | “Job will be boring/unglamorous” |
|  |  | Hospital issues | “Working in stretched system” |
| Logistics | Logistics | Accommodation | “Will need new accommodation, won’t be able to stay at home” |
|  |  | Transport | “I don’t have a car and would be unsure whether public transport would be appropriate - I live far out from major hospitals.” |
|  |  | Administrative | “Admin issues” |
|  | Support | Support outside of work | “Not being able to see my friends and family outside of my house will be difficult”; “Lack of peer and colleague support”; “Lack of university support” |
|  | Financial | Lack of pay | “I think if students are asked to join the workforce during a pandemic then they must be paid something.” |
|  |  | Financial difficulties | “Students are considered financially vulnerable for clinical trial purposes, similar ethical concerns exist around paid volunteer opportunities” |
| Education | University education | Educational disruption | “If we choose not to volunteer, we risk being under skilled compared to our contemporaries who do volunteer”; “Even further loss of study time” |
|  |  | Work-educational balance | “Volunteering can take students away from their focus on the degree”; “we have exams imminently and have been told if we choose to volunteer we are expected to still perform in exams, and the exams will still contribute towards deciles ... Many of us have had to choose between doing what we can to help and risking our education” |
|  | Professional education | Lack of training | “Inappropriate clinical training”; “Without proper training/preparation, harm may also be done to patients or healthcare staff supervising students” |
|  |  | Learning how to work | “The 'learning' time it takes to learn how the job works” |

#### Deciding responsibility

Two themes were relevant to deciding responsibility: pressure to volunteer and education. Students expressed feelings such as guilt, obligation and, even a sense of coercion. There were concerns that opportunities may not be promoted in a neutral way and that it should be students’ own choice to volunteer rather than due to pressure from external organizations (Table 2).

The theme of education was also relevant to deciding responsibility. Some students felt that volunteering was an opportunity to replace disrupted teaching opportunities, whereas others found there was a conflict between volunteering and studying for their medical degree, which was compounded by a perceived lack of training for volunteers (Table 2).

#### Deciding whether you have the skills or knowledge to help

Regarding skills or knowledge required, we identified concerns surrounding the themes of professional practice and safety. Students were concerned about their competency and questioned the usefulness of inexperienced medical students, as they might constitute a burden. These concerns were closely linked to safe practice – that patient care might be affected due to working outside of competency, especially if there was a lack of supervision or clarity regarding the professional role boundaries of new doctors.

#### Additional factors which influenced making a final decision to provide help

The theme of safety also played a key role in decision making. This included considerations of personal safety, the risk of contracting COVID-19 (particularly with inadequate PPE, or for students with pre-existing health issues), as well as the psychological impact and stress of working during a pandemic. Students were concerned about transmitting COVID-19 to others, the risk posed to vulnerable family members, or disruption to pre-existing caring duties.

Our qualitative analysis revealed a final barrier not accounted for by Latane and Darley’s theory: ‘logistics’. These were concerns of support outside of work, and difficulties with transport, accommodation, and administration.

## DISCUSSION

In this study, we identified that 82.7% of medical student respondents would be willing to volunteer; however, only 34.5% were volunteering at the time of the survey. While other studies have shown a similar association, they have not explored the reasons *why* this may be the case in depth ^17–19^. Studies in other fields have demonstrated an intention to behavior gap ^34^, which may explain part of the discrepancy. However, such a large difference suggests the presence of additional factors influencing students’ decisions to volunteer. Latane and Darley’s theory of prosocial behavior during an emergency outlines a five-step process influencing the decision-making process to help in emergency situations that we have used as a conceptual framework^126^. But how do our findings fit with what is already known?

Although McCrea and Murdoch-Eaton suggest medical students possess little awareness of the applicability of prosocial behavior to their role ^11^, our study shows this does not appear to be the case during COVID-19. The majority of students *did* recognize that COVID-19 was an abnormal situation, that it was an emergency, and that they may be responsible. This may be due, in part, to widespread media coverage, but is also likely a result of clinical placement suspension, and students witnessing the vast number of healthcare staff involved in responding to the pandemic. Time to reflect on a situation ^35^, as well as the presence of others providing help, has been shown to influence likelihood of a decision to help ^36^.

The gap between an intention to volunteer and volunteering in practice could be explained by the ways in which students decided their responsibility to volunteer during in the pandemic. This decision was complex, and influencing factors can be conceptualized as a balance between the interest of others and self-interest. Although 82.7% of students were willing to volunteer, prosocial behaviors can be influenced by others ^37^. This is shown in our cohort by a ‘pressure to volunteer’. Prosocial peer norms ^38^, and external influences (such as school and parents) have been shown to motivate prosocial behavior ^37^. Individuals are more likely to volunteer with “in-groups” (those close to them) rather than “out-groups” (those distant to them) ^39^. Thus, increased social distance between helpers and those being helped reduces prosocial behavior ^40^, as does social exclusion ^41^. For medical students, academic and clinical studies were interrupted – especially for those in later years used to high levels of patient contact – with many medical students returning home because of COVID-19. This may have increased the perceived social distance between patients, as well as between peers, shifting the balance of prosocial behavior towards self-interest.

Latane and Darley suggest that, after having assumed responsibility, people decide whether they have appropriate skills to help during emergencies ^12^. Similarly, we found that medical students make a series of ‘competency’ judgements regarding volunteering, which includes considerations as to how their level of ability relates to the proposed volunteering role and level of supervision, and how these factors might influence patient safety. There was an incremental increase in their confidence in the required skills and their willingness to perform more advanced roles as the year of the student increased, such that final year students were less willing to provide indirect care even though they had the skills to do so. Our data suggest that this decision depends not only on the skills required, but also on the *roles* offered and the year of the student. We posit that this is a domain which could result in a large decrease in prosocial behavior during COVID-19. Our qualitative data showed that students felt uncertain as to the professional role boundaries of volunteers; they perceived a lack of clarity regarding voluntary roles and were worried that they might be asked to perform tasks outside of their competence. Concerns surrounding roles could also reflect the variation of roles across the country ^42–46^. Clarity regarding roles and necessary skills could facilitate prosocial behavior amongst medical students during emergencies. Presenting students with more options involving a wider range of roles could also help, as having a number of options by which one could help has been shown to facilitate prosocial behavior ^47^.

Finally, in Latane and Darley’s theory, an individual weighs the above considerations and makes a final decision ^12^. We found that significantly fewer students who were intercalating (taking time out of medical school to complete an additional degree) volunteered. This lends weight to our observation about social distance, as intercalating students may be further removed from patients, healthcare professionals, and medical schools. We also showed that far more final-year students had volunteered than first-year students. This supports our arguments about role and skills alignment. Students in higher years are likely to possess a higher level of self-efficacy – the ability to overcome barriers to achieve a goal – which can influence prosocial behavior ^6,48^. Interestingly, in contrast to this finding, Burks and Kobus found that prosocial values decrease as students’ progress through training ^7^. We posit that the main reason for the difference between this study and Burks and Kobus’ research is that final year students had a clear interim postgraduate year one role that they could fulfil, which students in other years lacked. Regarding barriers to volunteering, 43.5% of respondents felt there was lack of information on volunteering opportunities available. ‘Logistics’ was the first of two areas that had not been explained in Latane and Darley’s theory of prosocial behavior during an emergency. Our data would indicate that – a volunteer must decide whether *logistically* they can volunteer. Previous literature surrounding the willingness of medical students to volunteer in a disaster relied on the assumption that in a crisis there would be an established framework and infrastructure for mobilizing medical students. However, this was not the case in the UK during the early part of 2020. Early narrative work from the USA has centered on development of volunteering infrastructure, including the establishment of voluntary task forces ^49^, with similar initiatives occurring in the UK ^50^.

The second key area that had not been adequately encapsulated by Latane and Darley’s theory concerned ‘safety’. Although considerations of safety may take place within decisions of responsibility, or of skills and knowledge, our data demonstrate that concerns regarding safety were an integral part of volunteering decision-making. We posit that medical students must also decide whether it is safe to volunteer – in the interests of themselves and others. We present our conceptual framework for medical student volunteering during COVID-19 in Figure 2.

**Figure 2:**
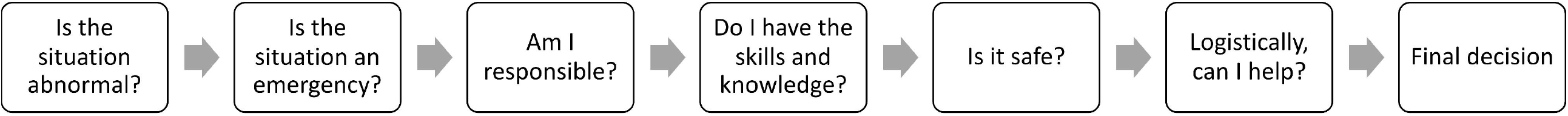
Conceptual framework for medical student prosocial behaviour during COVID-19.

Understanding the factors that influence prosocial behavior during COVID-19 can support future decision-making around the infrastructure and processes that must be put in place to effectively facilitate the mobilization of students during the current pandemic and in any future crises. Creating a comprehensive strategy for how to manage and implement volunteers is beyond the scope of this article. We have provided suggestions in Table 3 of how our conceptual framework can be used in educational strategies to facilitate medical student prosocial behavior during pandemics and disasters ^51^. These could be introduced using pre-existing frameworks for innovation, and could be used to develop a flexible structure that is organized at a local level with national oversight, which could allow for a rapid goal-orientated coordinated response ^52,53^. Developing this infrastructure is even more important in view of the ‘second wave’ of COVID-19 cases ^54,55^ in Europe ^56^, and high numbers of cases in the USA ^57^.

**Table 3:**
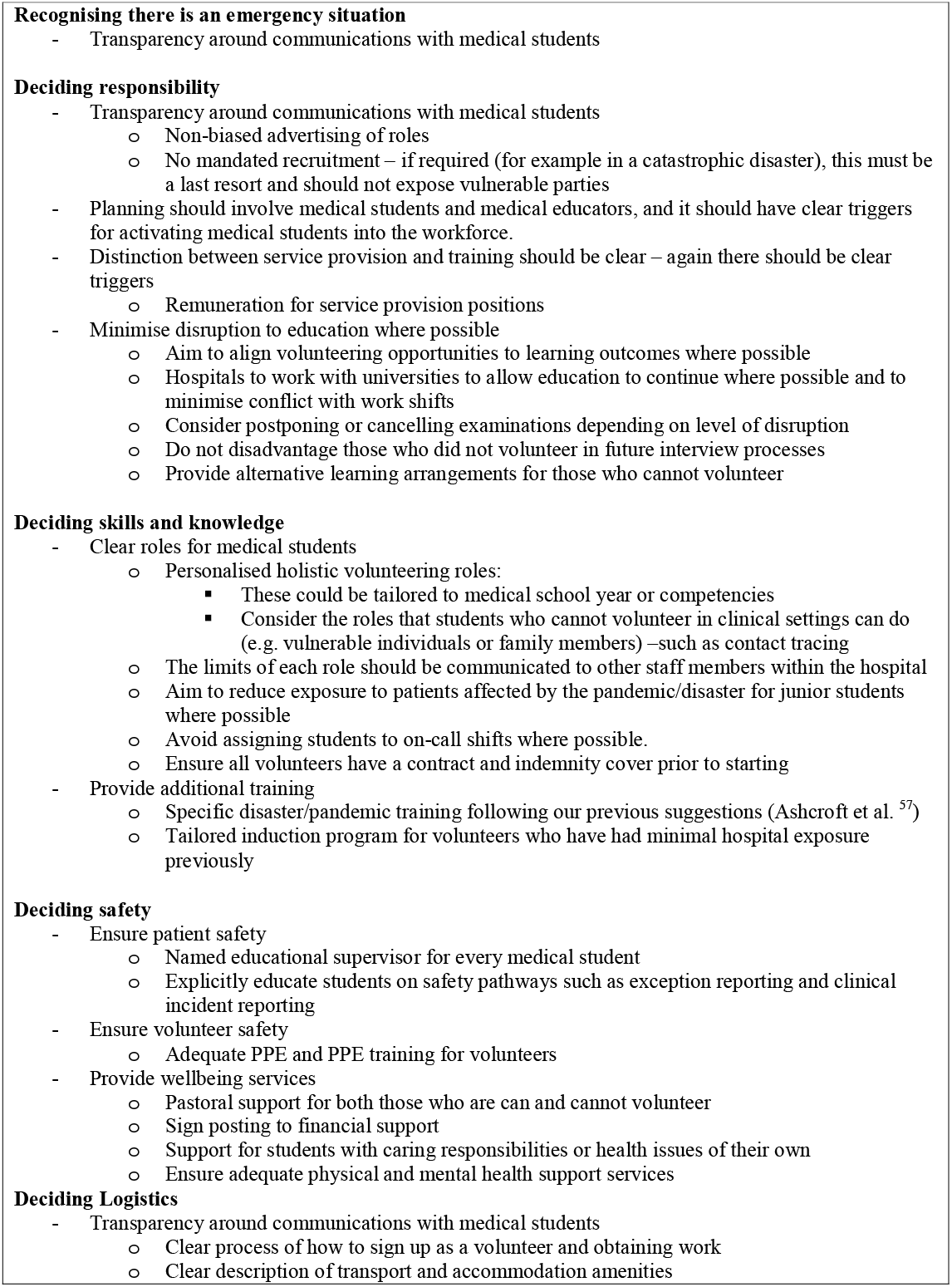
Suggestions to facilitate medical student prosocial behaviour during pandemics and disasters

## Limitations

To expedite survey distribution, we did not perform focus groups and cognitive interviews as part of our survey development, and this may have limited how participants interpretated the questions ^58^. However, medical students were involved throughout the survey development process. As we were unable to identify the survey response rate, our data may not be wholly representative of the whole UK cohort. We tried to mitigate this by distributing the survey through multiple channels. There is selection bias, as the types of medical student who opt to fill in the survey may be more willing to volunteer. Further, the transferability of our findings may be limited by the higher number of women who participated. Finally, this study was cross-sectional in nature, and could not determine whether students’ motivation to volunteer evolved as the context of the pandemic changed. Longitudinal research regarding the ways in which prosocial behaviors expression changes as emergency situations develop would be of benefit in future.

## CONCLUSIONS

This study demonstrates that Latane and Darley’s theory of prosocial behavior during an emergency can be applied to medical student volunteering during COVID-19. This study expands on existing theory through addition of the domains of *safety* and *logistics* in the decision-making process. We identified a number of modifiable barriers to prosocial behavior encountered by medical students during COVID-19 and provide suggestions of how our conceptual framework can be used within educational strategies to address such barriers. Optimizing the process of volunteering can aid workforce planning and healthcare provision, and may facilitate a volunteering process that is safer for students, staff, and patients.

## Data Availability

The data that support the findings of this study are available from the corresponding author upon reasonable request.

## AUTHOR DESCRIPTIONS

Matthew H V Byrne is an Academic Clinical Fellow in Urology at Oxford. He has a PGCert in medical education and has previously been awarded a MRes in Transplantation. He founded a national charity that facilitates medical student volunteers delivering talks at secondary schools.

James Ashcroft is an Academic Clinical Fellow in General Surgery in the East of England Deanery with an interest in early academic education and technical performance training. His educational work has explored cognitive interventions in technical skill (awarded MRes) and early surgical learning (awarded MSc).

Dr Laith Alexander is currently an Academic Foundation Doctor in Psychological Medicine & Psychiatry at Guy’s and St Thomas’ Foundation Trust with King’s College London. His interests lie in psychiatry and neuroscience. His PhD in neuroscience explored the role of the ventromedial prefrontal cortex in the regulation of emotion and cardiovascular function.

Dr Jonathan Wan is an Academic Foundation Doctor in Oncology at Guy’s and St Thomas’. He carried out his PhD in computational cancer diagnostics and is continuing his research as a Bioinformatics Engineer at Memorial Sloan Kettering Cancer Center. He is interested in medical and surgical oncology research and medical education.

Anna Harvey is a final year medical student at King’s College London and former Editorial Scholar at The BMJ.

Dr Nicholas Schindler is a pediatric registrar at Norfolk and Norwich University Hospitals NHS Foundation Trust, and a Tutor in Medical Education the University of Cambridge Institute of Continuing Education. His research interest is in methods of workplace-based training and assessment in pediatric education (awarded MSt). As Royal College of Pediatrics and Child Health trainee representative for Assessment and Examinations he is involved in the adaptation of assessments and postgraduate examinations for pediatricians during the COVID-19 pandemic.

Dr Megan Brown is a PhD student in medical education within the Health Professions Education Unit, Hull York Medical School. Her research interests include professional identity development, longitudinal integrated clerkships, empathy and the hidden curriculum.

Dr Cecilia Brassett is a Fellow at Magdalene College, Cambridge as well as their Joint Director of Studies in Medical Sciences, College Lecturer in Medicine, and Deputy Senior Tutor. She is also the University Clinical Anatomist. She is directly involved in the changes being made to medical student teaching during the COVID-19 pandemic.

## SUPPLEMENTARY FIGURES

Figure S 1: Respondent demographics. (a) Number of medical students from each medical school, divided by gender. (b) Total number of students, by gender. (c) Distribution of year groups of medical students divided by gender. (d) Pie chart indicating if students had already started as volunteers during the pandemic. (e) Proportion of students volunteering by year. The proportion of non-intercalating students volunteering at the time of the survey ranged from 14.6% among first-year students to 62.8% in sixth-year students

Figure S 2: Opinions on volunteering on a Likert scale.

Figure S 3: Roles students are willing to perform while volunteering grouped by year.

## REFERENCES

1. Siddique H. Final-year medical students graduate early to fight Covid-19. The Guardian. 2020.

2. Iacobucci G. Covid-19: medical schools are urged to fast-track final year students. BMJ. 2020;368:m1064. doi:10.1136/bmj.m1064

3. Medical Schools Council. STATEMENT OF EXPECTATION Medical Student Volunteers in the NHS.; 2020.

4. O’Byrne L, Gavin B, McNicholas F. Medical students and COVID-19: the need for pandemic preparedness. J Med Ethics. 2020:medethics-2020-106353. doi:10.1136/medethics-2020-106353

5. Arora A, Solomou G, Bandyopadhyay S, et al. Adjusting to Disrupted Assessments, Placements and Teaching (ADAPT): a snapshot of the early response by UK medical schools to COVID-19. medRxiv. August 2020:2020.07.29.20163907. doi:10.1101/2020.07.29.20163907

6. Baron RA, Byrne D, Branscombe NR. Mastering Social Psychology. Boston: Pearson Education, Inc; 2007.

7. Burks DJ, Kobus AM. The legacy of altruism in health care: The promotion of empathy, prosociality and humanism. Med Educ. 2012;46(3):317–325. doi:10.1111/j.1365-2923.2011.04159.x

8. Arnold EL, Blank LL, Race KEH, Cipparrone N. Can professionalism be measured? The development of a scale for use in the medical environment. Acad Med. 1998;73(10):1119–1121. doi:10.1097/00001888-199810000-00025

9. Leffel GM, Oakes Mueller RA, Ham SA, Karches KE, Curlin FA, Yoon JD. Project on the Good Physician: Further Evidence for the Validity of a Moral Intuitionist Model of Virtuous Caring. Teach Learn Med. 2018;30(3):303–316. doi:10.1080/10401334.2017.1414608

10. Patterson F, Zibarras L, Ashworth V. Situational judgement tests in medical education and training: Research, theory and practice: AMEE Guide No. 100. Med Teach. 2016;38(1):3–17. doi:10.3109/0142159X.2015.1072619

11. McCrea ML, Murdoch-Eaton D. How do undergraduate medical students perceive social accountability? Med Teach. 2014;36(10):867–875. doi:10.3109/0142159X.2014.916784

12. Latané B, Darley JM. The Unresponsive Bystander: Why Doesn’t He Help? New York: Appleton-Century Crofts; 1970.

13. Gouda P, Kirk A, Sweeney A-M, O’Donovan D. Attitudes of Medical Students Toward Volunteering in Emergency Situations. Disaster Med Public Health Prep. September 2019:1–4. doi:10.1017/dmp.2019.81

14. Patel VM, Dahl-Grove D. Disaster Preparedness Medical School Elective: Bridging the Gap Between Volunteer Eagerness and Readiness. Pediatr Emerg Care. 2018;34(7):492–496. doi:10.1097/PEC.0000000000000806

15. Rasmussen S, Sperling P, Poulsen MS, Emmersen J, Andersen S. Medical students for health-care staff shortages during the COVID-19 pandemic. Lancet. 2020;395(10234):e79–e80. doi:10.1016/S0140-6736(20)30923-5

16. Astorp MS, Sørensen GVB, Rasmussen S, Emmersen J, Erbs AW, Andersen S. Support for mobilising medical students to join the covid-19 pandemic emergency healthcare workforce: A cross-sectional questionnaire survey. BMJ Open. 2020;10(9). doi:10.1136/bmjopen-2020-039082

17. Yu N-Z, Li Z-J, Chong Y-M, et al. Chinese medical students’ interest in COVID-19 pandemic. World J Virol. 2020;9(3):38–46. doi:10.5501/wjv.v9.i3.38

18. Drexler R, Hambrecht JM, Oldhafer KJ. Involvement of Medical Students During the Coronavirus Disease 2019 Pandemic: A Cross-Sectional Survey Study. Cureus. 2020;12(8). doi:10.7759/cureus.10147

19. Pasquier P, Luft A, Gillard J, et al. How do we fight COVID-19? Military medical actions in the war against the COVID-19 pandemic in France. BMJ Mil Heal. August 2020:bmjmilitary-2020-001569. doi:10.1136/bmjmilitary-2020-001569

20. Patel J, Robbins T, Randeva H, et al. Rising to the challenge: Qualitative assessment of medical student perceptions responding to the COVID-19 pandemic. Clin Med (Northfield Il). October 2020:clinmed.2020-0219. doi:10.7861/clinmed.2020-0219

21. The General Medical Council. The State of Medical Education and Practice in the UK 2020.; 2020.

22. The General Medical Council. Medical School Annual Return 2017/2018.; 2018.

23. Cresswell J, Plano Clark V. Designing and Conducting Mixed Methods Research. Thousand Oaks, California: SAGE; 2007.

24. Brown MEL, Dueñas AN. A Medical Science Educator’s Guide to Selecting a Research Paradigm: Building a Basis for Better Research. Med Sci Educ. 2020;30(1):545–553. doi:10.1007/s40670-019-00898-9

25. Schifferdecker KE, Reed VA. Using mixed methods research in medical education: Basic guidelines for researchers. Med Educ. 2009;43(7):637–644. doi:10.1111/j.1365-2923.2009.03386.x

26. STROBE Initiative. STROBE Checklist for cross-sectional studies.

27. Byrne M, Ashcroft J, Alexander L, Wan J, Harvey A. 986 A Systematic Review of Medical Student Willingness to Volunteer and Preparedness for Pandemics and Disasters. Br J Surg. 2021;108(Supplement_2). doi:10.1093/bjs/znab134.185

28. Medical Schools Council. Medical schools.

29. R Core Team. R: A language and environment for statistical computing. 2017.

30. Braun V, Clarke V. Using thematic analysis in psychology. Qual Res Psychol. 2006;3(2):77–101. doi:10.1191/1478088706qp063oa

31. Varpio L, Paradis E, Uijtdehaage S, Young M. The Distinctions between Theory, Theoretical Framework, and Conceptual Framework. Acad Med. 2020;95(7):989–994. doi:10.1097/ACM.0000000000003075

32. Bowen GA. Grounded Theory and Sensitizing Concepts. Int J Qual Methods. 2006;5(3):12–23. doi:10.1177/160940690600500304

33. Patel R, Tarrant C, Bonas S, Yates J, Sandars J. The struggling student: a thematic analysis from the self-regulated learning perspective. Med Educ. 2015;49(4):417–426. doi:10.1111/medu.12651

34. Webb TL, Joseph J, Yardley L, Michie S. Using the Internet to Promote Health Behavior Change: A Systematic Review and Meta-analysis of the Impact of Theoretical Basis, Use of Behavior Change Techniques, and Mode of Delivery on Efficacy. J Med Internet Res. 2010;12(1):e4. doi:10.2196/jmir.1376

35. Darley JM, Batson CD. “From Jerusalem to Jericho”: A study of situational and dispositional variables in helping behavior. J Pers Soc Psychol. 1973;27(1):100–108. doi:10.1037/h0034449

36. Latané B, Darley JM. Group inhibition of bystander intervention in emergencies. J Pers Soc Psychol. 1968;10(3):215–221. doi:10.1037/h0026570

37. Lai FHY, Siu AMH, Shek DTL. Individual and Social Predictors of Prosocial Behavior among Chinese Adolescents in Hong Kong. Front Pediatr. 2015;3. doi:10.3389/fped.2015.00039

38. Choukas-Bradley S, Giletta M, Cohen GL, Prinstein MJ. Peer Influence, Peer Status, and Prosocial Behavior: An Experimental Investigation of Peer Socialization of Adolescents’ Intentions to Volunteer. J Youth Adolesc. 2015;44(12):2197–2210. doi:10.1007/s10964-015-0373-2

39. DiDonato TE, Ullrich J, Krueger JI. Social Perception as Induction and Inference: An Integrative Model of Intergroup Differentiation, Ingroup Favoritism, and Differential Accuracy. J Pers Soc Psychol. 2011;100(1):66–83. doi:10.1037/a0021051

40. Krueger JI, Ullrich J, Chen LJ. Expectations and decisions in the Volunteer’s dilemma: Effects of social distance and social projection. Front Psychol. 2016;7(DEC). doi:10.3389/fpsyg.2016.01909

41. swenge JM, Ciarocco NJ, Baumeister RF, DeWall CN, Bartels JM. Social exclusion decreases prosocial behavior. J Pers Soc Psychol. 2007;92(1):56–66. doi:10.1037/0022-3514.92.1.56

42. Youssef S, Zaidi S, Shrestha S, Varghese C, Rajagopalan S. First impressions of the foundation interim year 1 postings: positives, pitfalls, and perils. Med Educ Online. 2020;25(1):1785116. doi:10.1080/10872981.2020.1785116

43. UK Foundation Programme Office. Impact of COVID-19 on the UK Foundation Programme 2020. 2020.

44. Lee J. How medical students are responding to Covid-19. The Independent.

45. Kinder F, Harvey A. Covid-19: the medical students responding to the pandemic. BMJ. 2020;369:m2160. doi:10.1136/bmj.m2160

46. Thomson E, Lovegrove S. ‘Let us Help’—Why senior medical students are the next step in battling the COVID□19 Pandemic. Int J Clin Pract. 2020;74(8). doi:10.1111/ijcp.13516

47. Wang Z, Jusup M, Shi L, Lee JH, Iwasa Y, Boccaletti S. Exploiting a cognitive bias promotes cooperation in social dilemma experiments. Nat Commun. 2018;9(1). doi:10.1038/s41467-018-05259-5

48. Almeida L, Kashdan TB, Coelho R, Albino-Teixeira A, Soares-da-Silva P. Healthy subjects volunteering for phase I studies: Influence of curiosity, exploratory tendencies and perceived self-efficacy. Int J Clin Pharmacol Ther. 2008;46(3):109–118. doi:10.5414/CPP46109

49. Soled D, Goel S, Barry D, et al. Medical Student Mobilization During A Crisis: Lessons From A COVID-19 Medical Student Response Team. Acad Med. April 2020. doi:http://dx.doi.org/10.1097/ACM.0000000000003401

50. Medical Schools Council. Student volunteering.

51. Ashcroft J, Byrne MHV, Brennan PA, Davies RJ, Davies RJ. Preparing medical students for a pandemic: A systematic review of student disaster training programmes. Postgrad Med J. 2020;0:1–12. doi:10.1136/postgradmedj-2020-137906

52. Sheather J, Jobanputra K, Schopper D, et al. A Médecins Sans Frontières Ethics Framework for Humanitarian Innovation. PLoS Med. 2016;13(9). doi:10.1371/journal.pmed.1002111

53. Whittaker J, McLennan B, Handmer J. A review of informal volunteerism in emergencies and disasters: Definition, opportunities and challenges. Int J Disaster Risk Reduct. 2015;13:358–368. doi:10.1016/j.ijdrr.2015.07.010

54. Wise J. Covid-19: Risk of second wave is very real, say researchers. BMJ. 2020;369:m2294. doi:10.1136/bmj.m2294

55. Xu S, Li Y. Beware of the second wave of COVID-19. Lancet. 2020;395(10233):1321–1322. doi:10.1016/S0140-6736(20)30845-X

56. Willsher K, Henley J. Covid in Europe: second wave gathers pace across continent. The Guardian.

57. Center for Disease Control and Prevention. Cases in the US.

58. Artino AR, La Rochelle JS, Dezee KJ, Gehlbach H. Developing questionnaires for educational research: AMEE Guide No. 87. Med Teach. 2014;36(6):463–474. doi:10.3109/0142159X.2014.889814

