## Supplementary material for "Examining Medical Student Volunteering During The COVID-19 Pandemic As A Prosocial Behavior During An Emergency": Table S 1

Table S 1: Volunteering role. Total percentage is >100% as 59 students held multiple roles during the pandemic. Allied healthcare professionals/technicians included roles such as occupational therapists, physiotherapists, radiographers, and clinical physiologists; caring roles were either in hospital or in care homes; administrative roles included roles such as ward clerk, secretary and receptionist, but also 111 callers; and healthcare scientists included those who were actively involved in clinical and laboratory research, as well as those working as biomedical scientists in hospital laboratories.

| Role | Role | |
| --- | --- | --- |
|  | n= | % |
| Healthcare assistant | 172 | 44.3 |
| Allied healthcare professional/technician | 29 | 7.5 |
| Carer/care assistant | 4 | 1 |
| Healthcare scientist | 15 | 3.9 |
| Pharmacy assistant | 6 | 1.5 |
| First responder/first aider | 2 | 0.5 |
| Pharmacist | 6 | 1.5 |
| Administrative role | 71 | 18.3 |
| Hospital volunteer | 2 | 0.5 |
| Paramedic/emergency medical technician | 4 | 1 |
| Nurse | 4 | 1 |
| Phlebotomist | 5 | 1.3 |
| Physician's assistant | 55 | 14.2 |
| Dentist | 2 | 0.5 |
| Doctor (interim foundation year one or equivalent) | 77 | 19.8 |
