## Supplementary figures and images for "Examining Medical Student Volunteering During The COVID-19 Pandemic As A Prosocial Behavior During An Emergency"

### Figure S 1

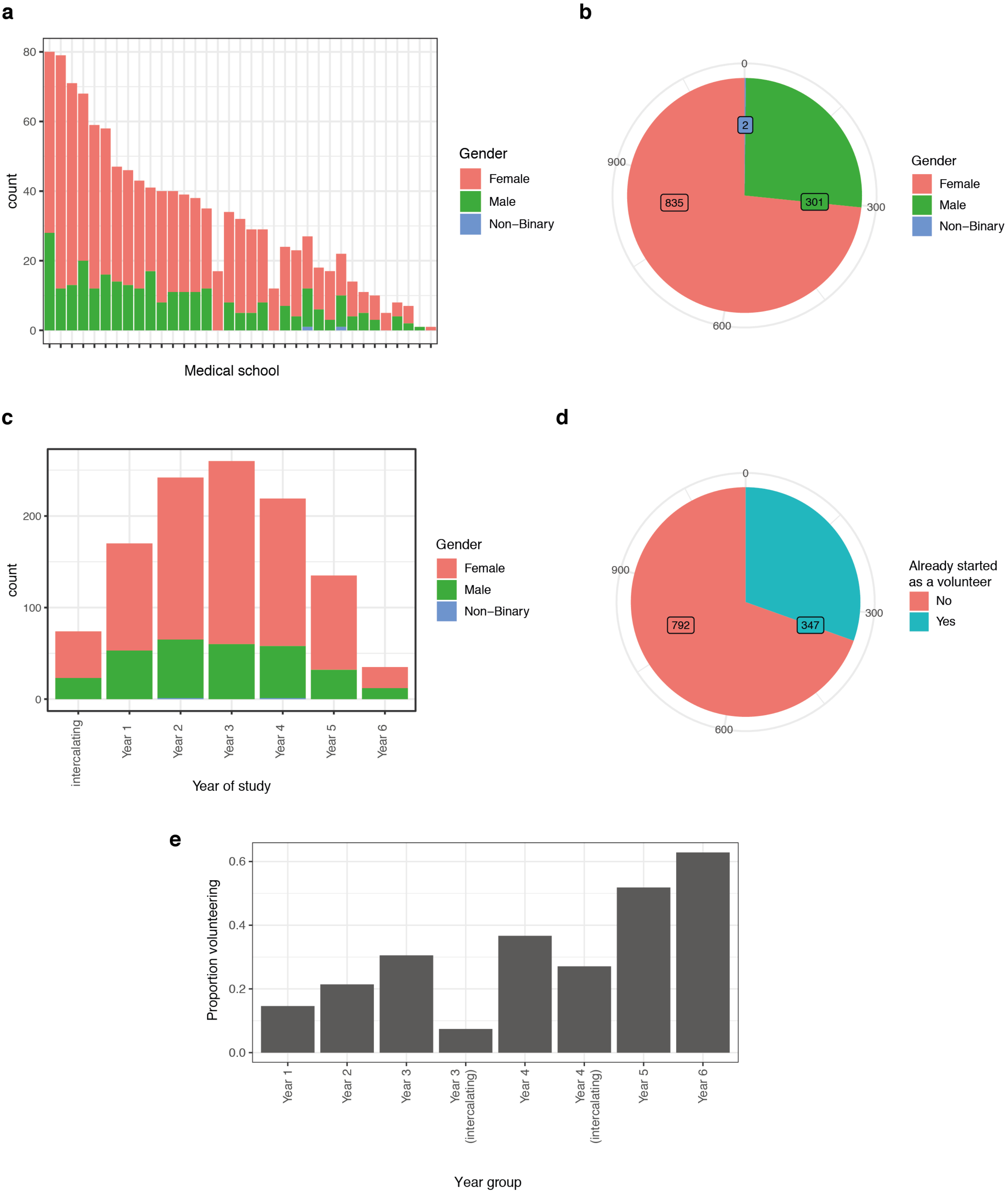

### Figure S 2

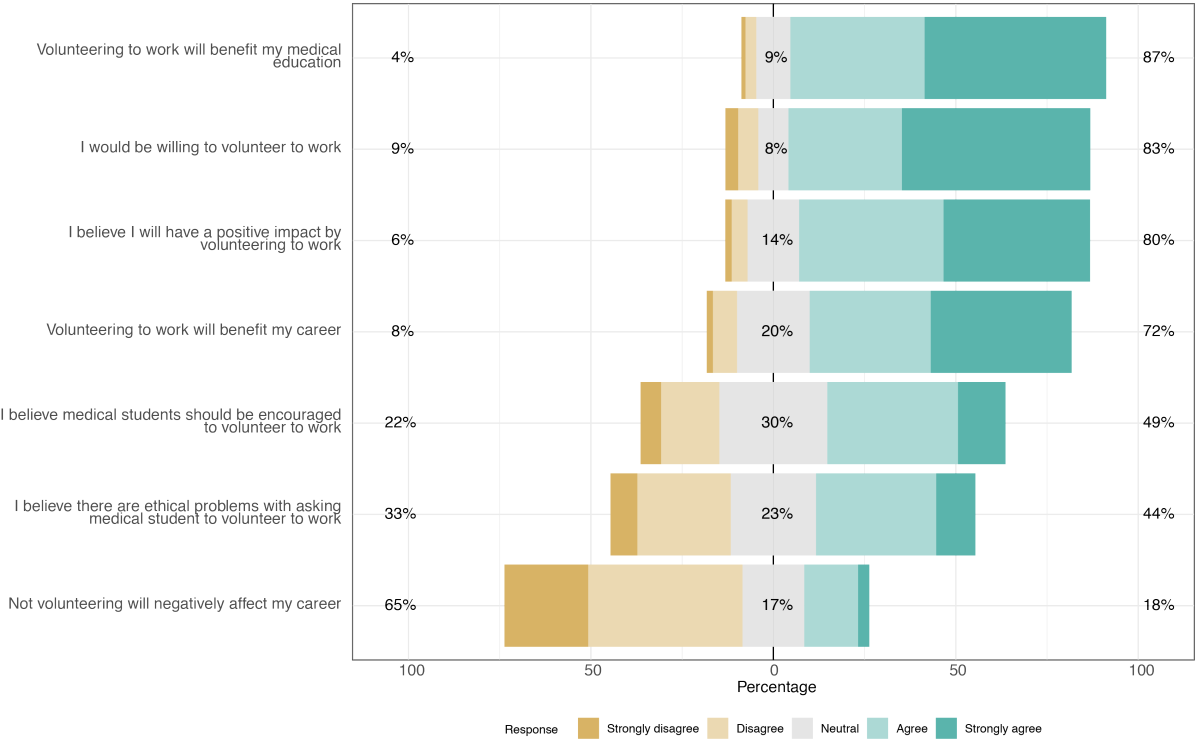

### Figure S 3

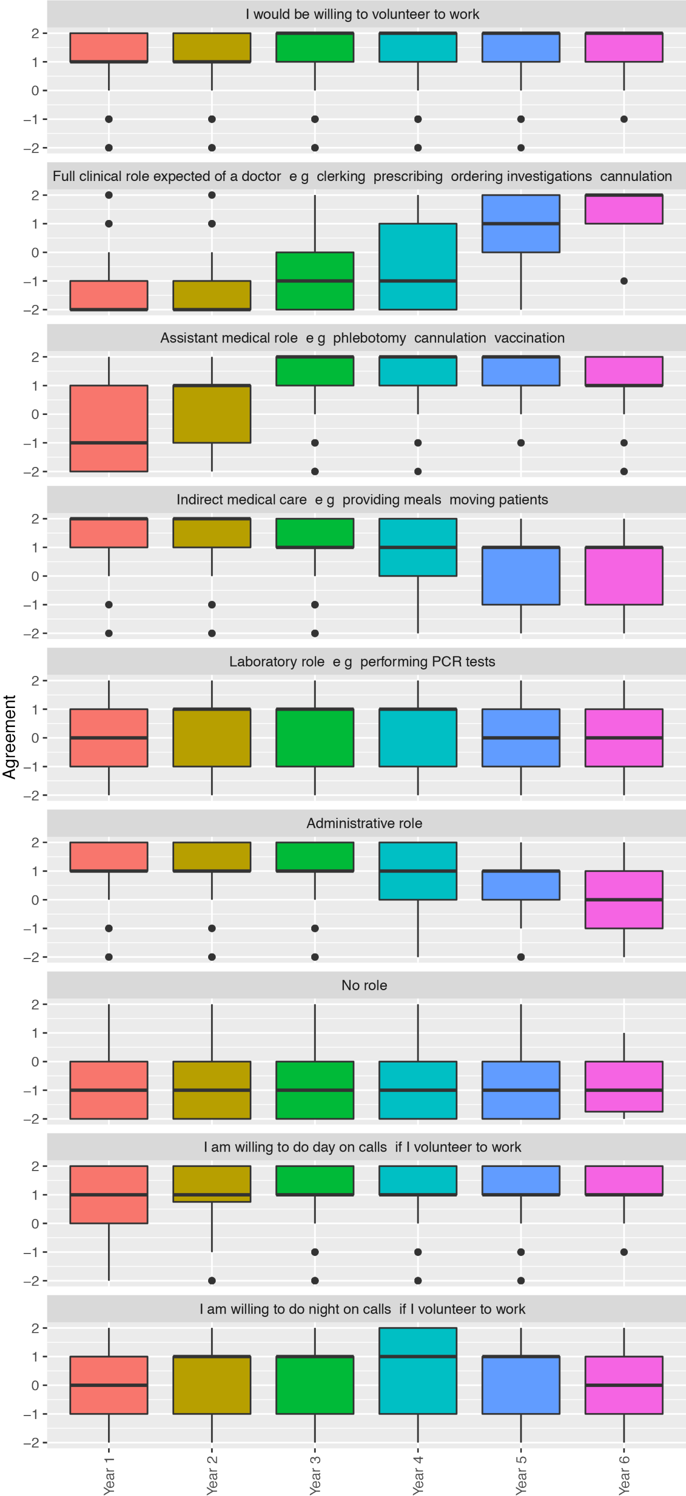

### Table S 2

*Table S 2: Multiple linear regression of predictors of willingness to volunteer.*

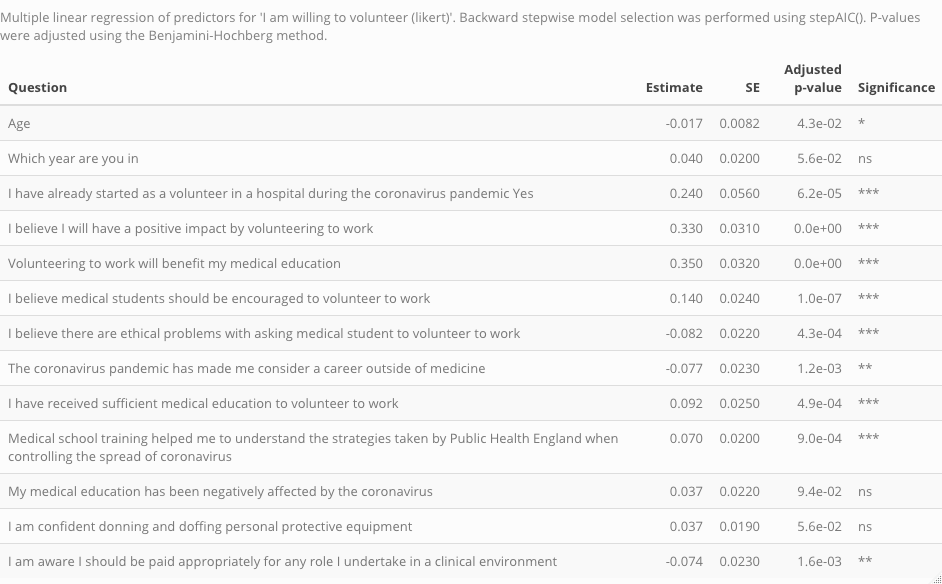
